# Personalized Synthetic Electrocardiograms with Outcomes

**DOI:** 10.1101/2025.08.01.25332716

**Authors:** Jonas L. Isaksen, Vajira Thambawita, Malene Nørregaard, Christina Ellervik, Torben Hansen, Allan Linneberg, Dominik Linz, Claus Graff, Michael A. Riegler, Jørgen K. Kanters

## Abstract

**Background:** Synthetic data can be the solution to privacy requirements, can enrich datasets limited by underrepresentation of certain subgroups/minorities, combat data shortage, and reduce annotation costs, to facilitate the development of data-hungry machine-learning applications. An important limitation of current synthetic data is the missing link to patient characteristics and outcomes. This metadata is essential for synthetic data to be useful for solving real-world problems, and we aimed to generate novel synthetic data with clinical characteristics and outcomes.

**Methods:** We designed a novel generative method for generating 1:1 personalized synthetic electrocardiograms (ECGs) with associated patient characteristics and outcomes. The architecture of the model is a U-net neural network, which creates patient-specific ECGs to allow attachment of patient characteristics, comorbidities, and outcomes. We developed the model on the General Suburban Population Study and the Lolland-Falster Study cohorts and validated the model on the Danish Inter99 cohort. We compared original and synthetic ECGs usings Bland-Altman plots, by comparing associations with sex, age, and body mass index using linear models, and by comparing associations with all-cause mortality using Cox models. We defined that at most 5% of synthetic ECGs should have their paired original ECG as nearest neighbor in Euclidean space.

**Results:** We generated 6,612 novel, personalized electrocardiograms. Although Bland-Altman plots showed a high level of agreement between synthetic and original ECGs, only 3.7% of synthetic ECGs had their original paired ECG as their nearest neighbor. Synthetic ECGs had preserved relations between heart rate, R-wave amplitude, and T-wave amplitude and age, sex, and body mass index. Corrected QT interval was about 9.5 ms shorter in men compared to women in both the original and synthetic cohorts. The associations between PR interval and clinical characteristics were attenuated in the synthetic cohort. Heart rate and corrected QT interval were each associated with increased mortality with similar hazard ratios in the original and synthetic populations.

**Conclusions:** We demonstrated the ability of a novel neural network method to generate personalized synthetic ECGs with preserved associations with many patient characteristics and with all-cause mortality. This method may facilitate data sharing for the development of better risk prediction models.

## Introduction

Reasonable privacy restrictions set forth by the European General Data Protection Regulation (GDPR) and the American Health Insurance Portability and Accountability Act (HIPAA) limit the sharing of private health data. Generating synthetic data based on real, clinical data is an established method to comply with privacy regulations.^1,2^ Synthetic data is artificially crafted data that can also help to enrich datasets with underrepresentation of certain patient groups, overcome data shortage, and reduce annotation costs.^3^ Use of synthetic data over real data has already improved calcium imaging,^4^ X-ray image analysis,^5^ DNA sequencing,^6^ and even earthquake predictions,^7^ and the European Union has allocated significant funds to the research in medical synthetic data, acknowledging its vital role in advancing healthcare and medical technologies.^8^

Medical time series, particularly the electrocardiogram (ECG), is a huge area of research with classical and machine learning methods alike. Seminal work has demonstrated that machine learning methods may someday interpret ECGs more accurately than medical doctors.^9^

Synthetic data generation can already generate unlimited, realistic ECGs, which, as a whole, retain the same characteristics as a desired population.^10^ However, these methods create only ECGs with no metadata. Most medical data, including ECGs, are useless without a link to metadata, including individual-level labels, patient characteristics, and outcomes. To overcome this issue, methods for personalized data synthesis are needed to create synthetic ECGs for a specific patient or patient group with well-defined characteristics and outcomes.

We aimed to develop and validate a generative neural network to create novel personalized ECGs with similar – but not identical – associations with patients characteristics and outcomes.

## Methods

### Populations

The training cohort consisted of 24,328 10-second 12-lead ECGs from the Danish General Suburban Population Study (GESUS) and the Lolland-Falster Study (LOFUS). The characteristics of these studies have been described in detail elsewhere.^11–13^ In short, for GESUS, residents aged 20 years and older in the Næstved municipality 70 km south of Copenhagen, Denmark were invited to participate. For LOFUS, residents aged 18 years and older were invited from the islands of Lolland and Falster to participate between 2016-2020.

The testing cohort consisted of 6,612 ECGs from participants in the baseline health examination of the Danish Inter99 cohort study. These participants were also invited based on Danish registers. The details of this study have been published elsewhere.^14^

All ECGs represented unique individuals. Participants in all cohorts provided written informed consent. The study complies with the Declaration of Helsinki. The studies were approved by institutional review boards and the Danish Data Protection Agency.

### Electrocardiogram preprocessing and selection

All standard 12-lead, 10-second ECGs were recorded using GE electrocardiographs (Wauwatosa, WI) at 500 Hz and re-analyzed using version 2.43 of the 12SL algorithm (GE Healthcare, Wauwatosa, WI).^15^ The personalized synthetic ECGs were uploaded and analyzed using the same algorithm. From original and synthetic ECGs we obtained the heart rate, QT interval, QTcF interval (corrected by the method of Fridericia^16^), QRS duration, PR interval, P-wave duration and P, R, and T amplitudes in leads II, V1, and V5. We defined the transition zone as happening in V1-V2, V3, V4, or V5-V6, as done previously.^17^

We included ECGs with sinus rhythm (12SL statements: Sinus rhythm, sinus bradycardia, normal sinus rhythm, sinus tachycardia, and marked sinus bradycardia)^15^ from participants aged 18 years or above. Pathological findings were allowed if the rhythm was sinus rhythm. We excluded ECGs that were deemed noisy based on semi-automatic assessment prior to this work.

Out of 8687 electronic ECGs in the GESUS cohort, we included 8496 ECGs (97.8%). 12 ECGs were noisy and 181 were not in sinus rhythm (2 ECGs were both noisy and not in sinus rhythm). From the LOFUS study, we included 15,832 of 18,997 ECGs (83.3%). 2610 ECGs (13.7%) were from people under the age of 18 years, 490 ECGs were not in sinus rhythm, and 89 ECGs were deemed noisy. From the Inter99 study, we included 6612 of 6671 (99.1%). 10 ECGs were deemed noisy and 49 were not in sinus rhythm.

We created one synthetic ECG for each original ECG. We added noise to each sample of each original ECG as a random number between 0 and 0.18 mV (uniform distribution) and supplied the noisy ECG to the model as input. The target output was the original, noise-free ECG, and the penalty (loss) was the point-by-point mean squared error. Because four of the twelve leads (III, aVR, aVL, aVF) are linearly dependent on leads I and II, we used an 8-lead ECG (leads I, II, V1-V6) and eventually calculated the remaining four leads (see the **Supplementary Material** for details).^18^

### Model

We trained a generative neural network supplied with an ECG plus noise to restore a noise-free ECG. The output was the personalized synthetic ECG. The model is of U-net architecture, based on our previous work,^10^ whereby the noisy ECG is first compressed in six steps (encoded) and then expanded in another 6 steps (decoded).^19^ The expansion stages receive information from the previous step as well as from the compression steps through skip connections (also termed residual connections). This architecture allows the model to synthesize personalized noise-free ECGs with small imperfections making them deviate slightly from the original ECGs.

During training, the neural network was rewarded if the mean squared error (MSE) between the original, noise-free ECG and the output (denoised ECG) was small. Notably, the MSE as a loss function takes the place of the discriminator in a classical Generative Adversarial network (GAN) setup, leaving only the generator to be trained. This loss function ensures – to a higher degree than a discriminator – that the phenotypes of the patient-specific time series are preserved in the generated synthetic time series, because a discriminator would learn the population and not the individuals.^10^

### Statistics

We used Bland-Altman plots to assess that individual ECG characteristics were preserved in the synthetic ECGs compared to the original paired ECGs. The Bland-Altman plots were supplemented with 95% confidence intervals for differences in ECG markers between the original and synthetic populations.

We tested that the relationships between ECG markers and sex, age, and body mass index, respectively, were preserved in the synthetic ECGs relative to the original ECGs using linear models.

We also assessed whether or not the relationship between ECG markers from synthetic ECGs and mortality was similar to the relationship between original ECG markers and mortality. We used Cox regression models to test the association between each ECG marker and all-cause mortality in the original and synthetic populations, respectively.

To ensure that synthetic ECGs were distinctly different from the original ECGs, we defined a 5% identification limit. We created a normalized multidimensional Euclidean space of ECG markers, i.e. we normalized QTcF interval, QRS duration, PR interval, R peak amplitude, and T peak amplitude for original and synthetic ECGs to zero mean and unit variance. Then, for each synthetic ECG, we found the nearest original ECG neighbor in the Euclidean space. We then counted for each synthetic ECG, if the nearest neighbor was the matching original ECG, or if it was not. We defined that the ECGs were sufficiently separated if the nearest neighbor among the original ECGs was the ECG used for synthesis in less than 5% of cases. **Supplementary Figure S1** visualizes the concept of the nearest neighbor.

Statistical analyses were conducted with R (version 4.1.2, R Foundation for Statistical Computing, Vienna, Austria). The model was trained in Python version 3.10 and implemented using PyTorch version 2.0.0+cu117.

## Results

The generative model converged using raw ECG waveforms from the baseline visits of the GESUS and LOFUS cohorts (n=24,328, 54% women, median age 57.6 years).

We generated 6,612 synthetic ECGs, one from each included participant of the Inter99 cohort. These participants were on median 45.1 years of age and 51% were women (**Table 1**).

**Table 1.**
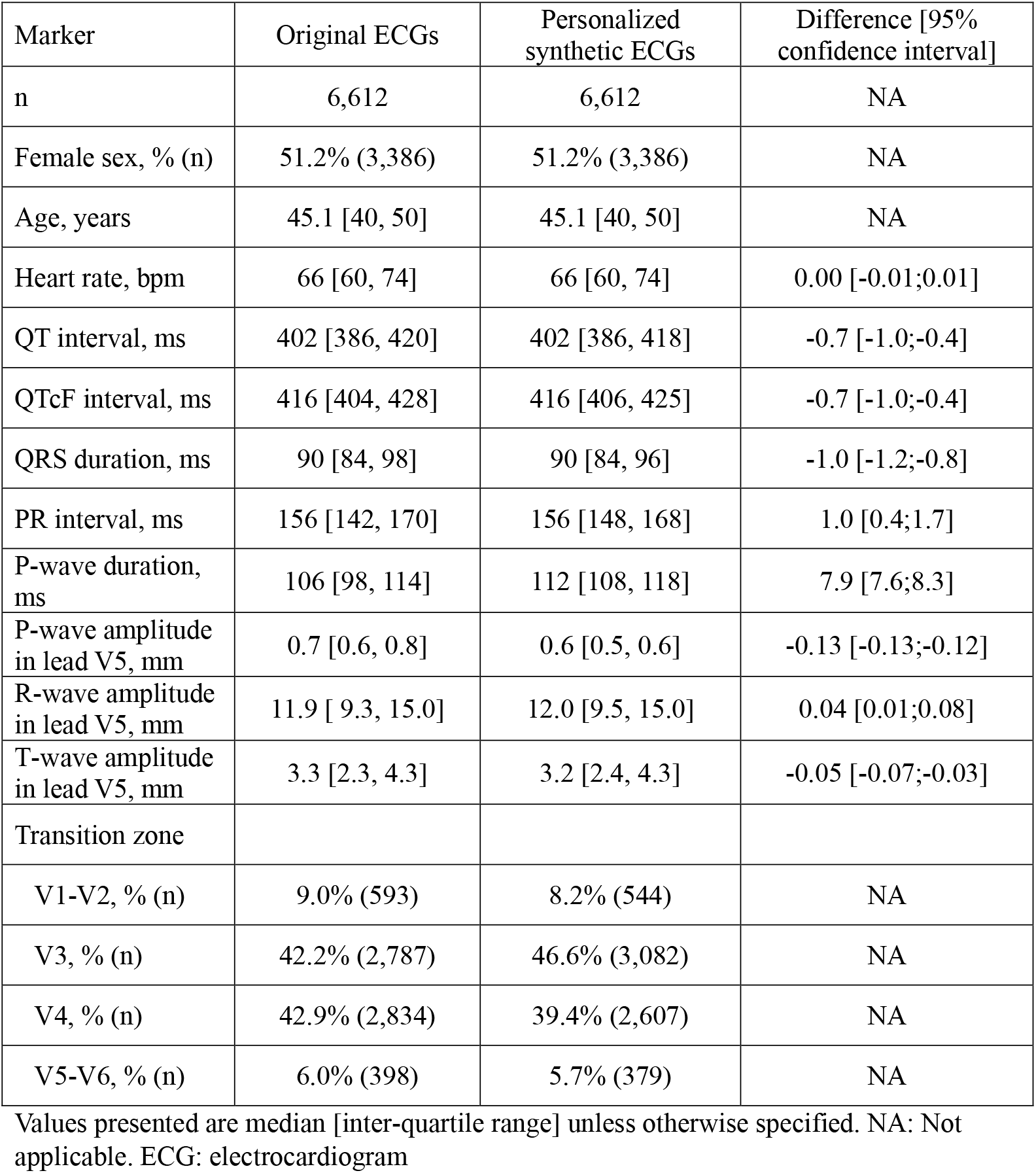
Characteristics of the original population and of recreated, personalized ECGs.

| Marker | Original ECGs | Personalized synthetic ECGs | Difference [95% confidence interval] |
| --- | --- | --- | --- |
| n | 6,612 | 6,612 | NA |
| Female sex, % (n) | 51.2% (3,386) | 51.2% (3,386) | NA |
| Age, years | 45.1 [40, 50] | 45.1 [40, 50] | NA |
| Heart rate, bpm | 66 [60, 74] | 66 [60, 74] | 0.00 [-0.01;0.01] |
| QT interval, ms | 402 [386, 420] | 402 [386, 418] | -0.7 [-1.0;-0.4] |
| QTcF interval, ms | 416 [404, 428] | 416 [406, 425] | -0.7 [-1.0;-0.4] |
| QRS duration, ms | 90 [84, 98] | 90 [84, 96] | -1.0 [-1.2;-0.8] |
| PR interval, ms | 156 [142, 170] | 156 [148, 168] | 1.0 [0.4;1.7] |
| P-wave duration, ms | 106 [98, 114] | 112 [108, 118] | 7.9 [7.6;8.3] |
| P-wave amplitude in lead V5, mm | 0.7 [0.6, 0.8] | 0.6 [0.5, 0.6] | -0.13 [-0.13;-0.12] |
| R-wave amplitude in lead V5, mm | 11.9 [ 9.3, 15.0] | 12.0 [9.5, 15.0] | 0.04 [0.01;0.08] |
| T-wave amplitude in lead V5, mm | 3.3 [2.3, 4.3] | 3.2 [2.4, 4.3] | -0.05 [-0.07;-0.03] |
| Transition zone |  |  |  |
| V1-V2, % (n) | 9.0% (593) | 8.2% (544) | NA |
| V3, % (n) | 42.2% (2,787) | 46.6% (3,082) | NA |
| V4, % (n) | 42.9% (2,834) | 39.4% (2,607) | NA |
| V5-V6, % (n) | 6.0% (398) | 5.7% (379) | NA |

Values presented are median [inter-quartile range] unless otherwise specified. NA: Not applicable. ECG: electrocardiogram

**Table 1** shows key ECG markers from the original and synthetic ECGs. As expected, heart rates were the same. The differences between global ECG markers for original and synthetic ECGs were at most 1 ms for the QTcF interval, QRS duration, and PR interval, but synthetic ECGs on average had an 8 ms longer P-wave duration in lead V5. In lead V5, P waves were 0.1 mm lower, R-wave amplitudes were 0.04 mm higher, and T-wave amplitudes were 0.05 mm smaller in synthetic ECGs compared to original ECGs.

Despite the similarities, we found that only 3.7% of ECGs (limit: 5%) were a nearest-neighbor match with their original, paired ECG (**Supplementary Figure S1**). Consequently, >96% of synthetic ECGs are more similar with another real ECG than with their source ECG used for generation measured by the ECG markers, suggesting that the synthetic ECGs are in fact novel tracings and not just copies of their source ECG.

Bland-Altman plots for key ECG markers are supplied as **Figure 1**. All Bland-Altman plots display the differences in means as described, but very little dependency on marker value were found except for the PR interval.

**Figure 1.**
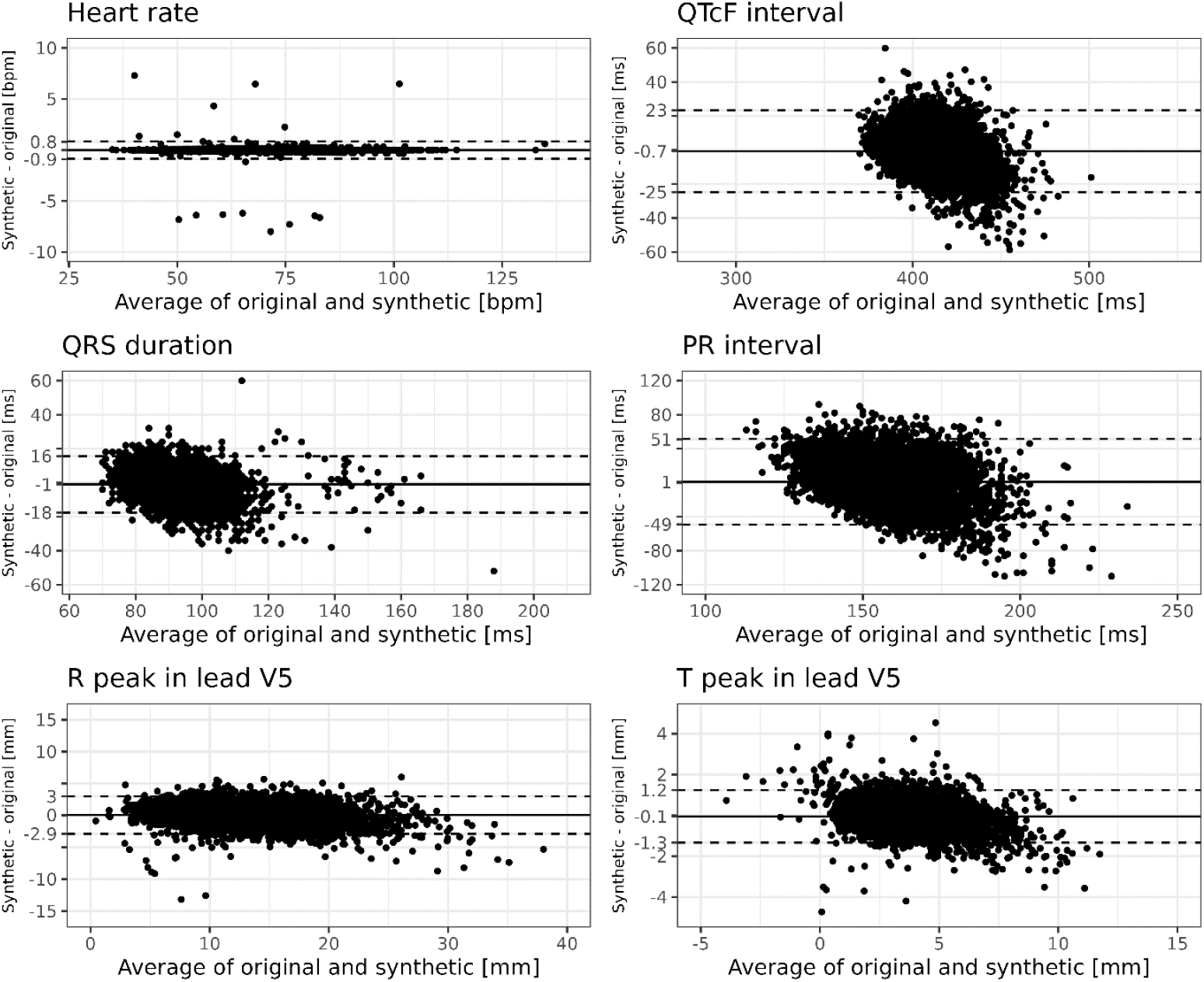
Bland-Altman plots for key electrocardiographic markers.

In addition to a high similarity between the synthetic and original ECGs in terms of clinically used ECG markers, also the associations between the ECGs markers and patient characteristics were similar for original and personalized synthetic ECGs. For age (**Figure 2**) we found the same associations with ECG markers in the original population and the synthetic population, except for the PR interval. The same was the case for the association between ECG markers and sex (**Figure 3**), with the exception of the QRS duration, for which the strength of the association was mildly reduced.

**Figure 2.**
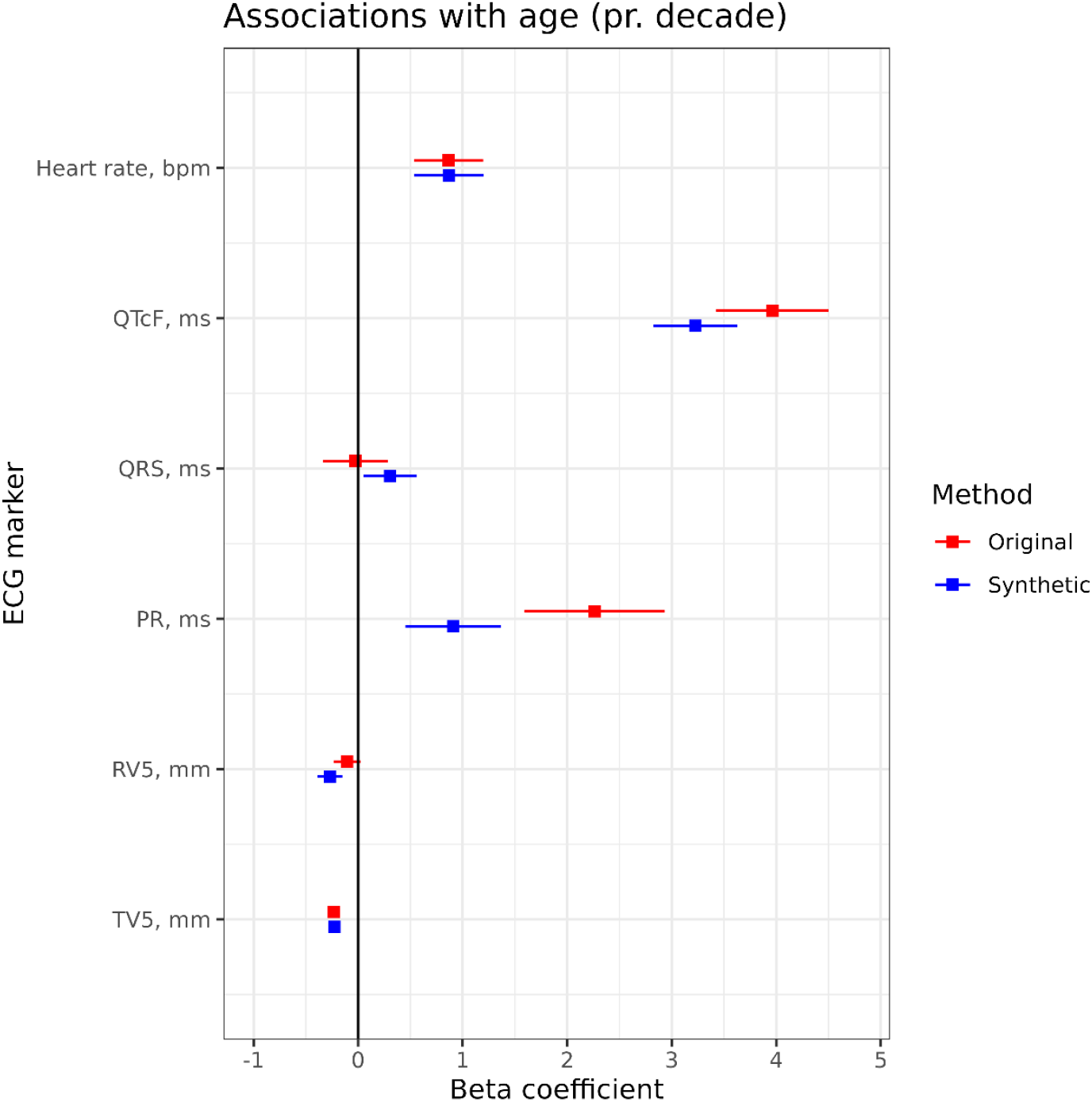
Associations between electrocardiogram (ECG) markers and age. We found significant and similar associations between age and heart rate, QT interval, PR interval, R-wave amplitude in lead V5, and T-wave amplitude in lead V5 in original and generated ECGs. We found no significant association between QRS duration and age in either the original population or the generated population.

**Figure 3.**
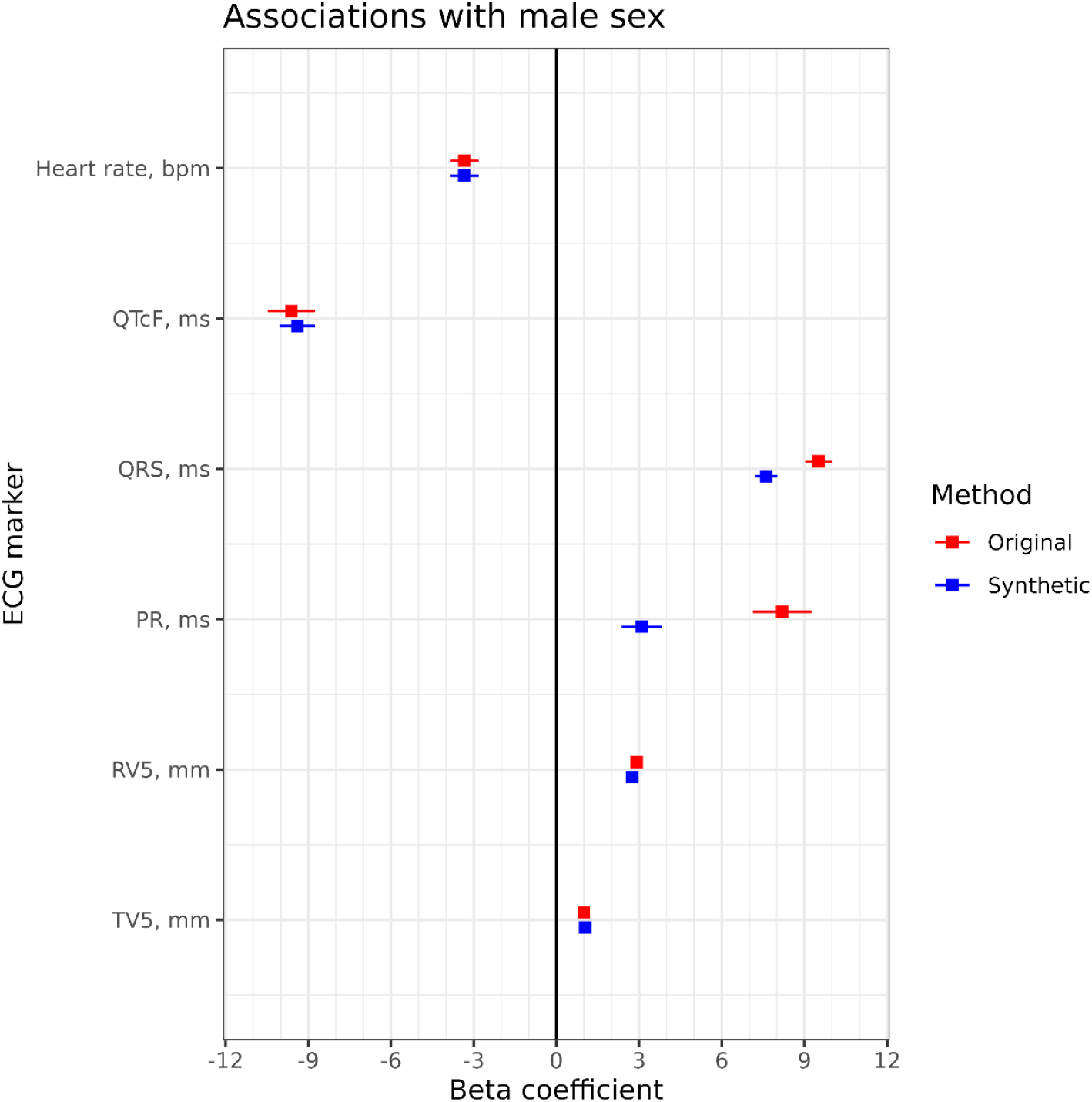
Associations between electrocardiogram (ECG) markers and sex. We found significant and similar associations between age and heart rate, QT interval, QRS duration, R-wave amplitude in lead V5, and T-wave amplitude in lead V5 in original and generated ECGs. The association between PR interval and sex was slightly attenuated in the generated ECGs compared with the original ECGs.

The associations between ECG markers and BMI were preserved for heart rate, R-wave amplitude, and T-wave amplitude, and mildly reduced for QTcF interval and PR interval (**Figure 4**).

**Figure 4.**
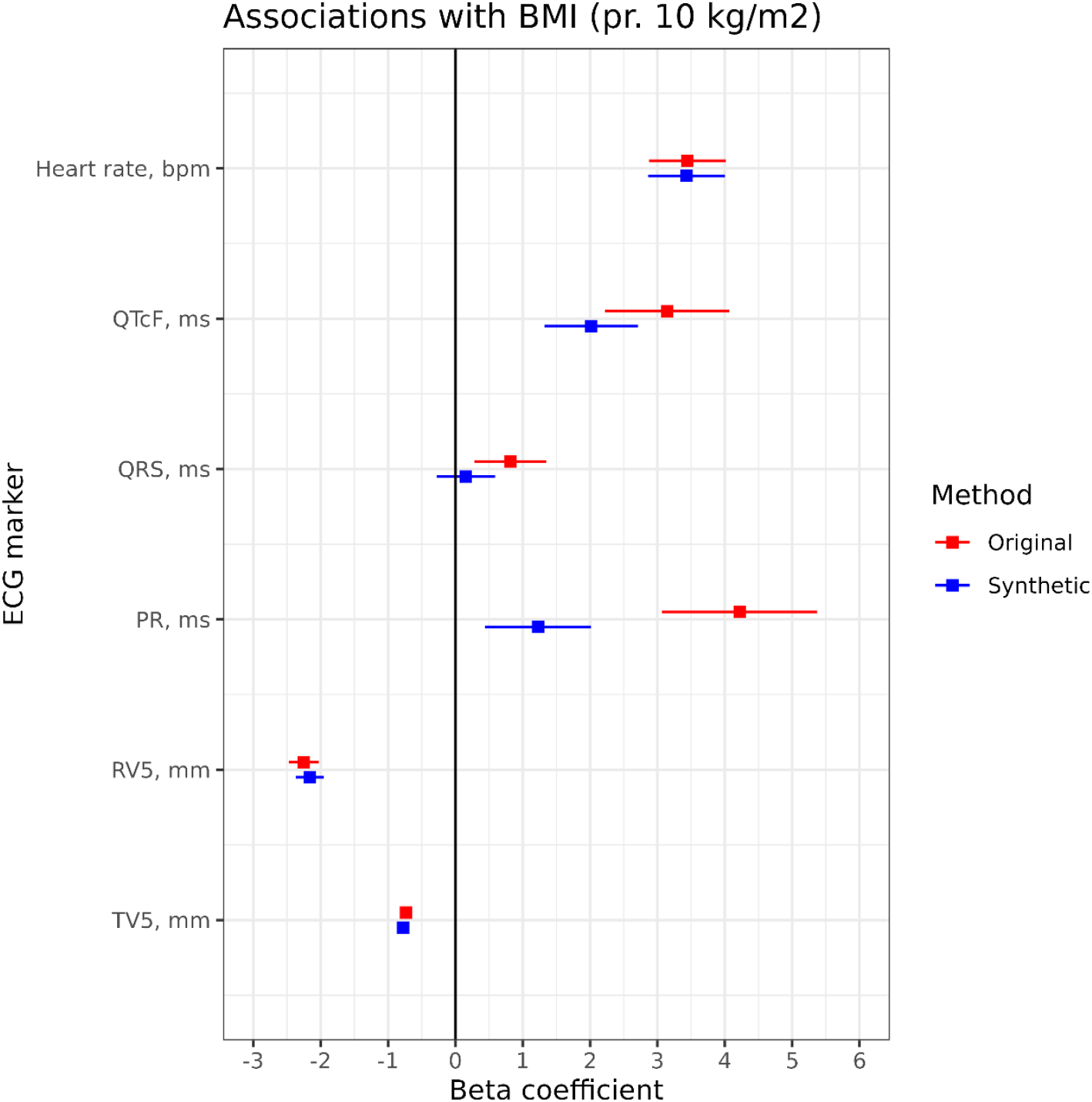
Associations between electrocardiogram (ECG) markers and body mass index (BMI). Associations were similar in the recreated ECGs and in the original ECGs for heart rate and R-wave and T-wave amplitudes. Associations were identified at a lower strength for the QTcF interval and the PR interval, and not for the QRS duration.

Importantly, we found that the association between ECG markers and all-cause mortality was similar between original and synthetic ECGs (**Figure 5**). Thus, for this hard clinical outcome the predictive value of the original ECGs is preserved in the synthetic ECGs

**Figure 5.**
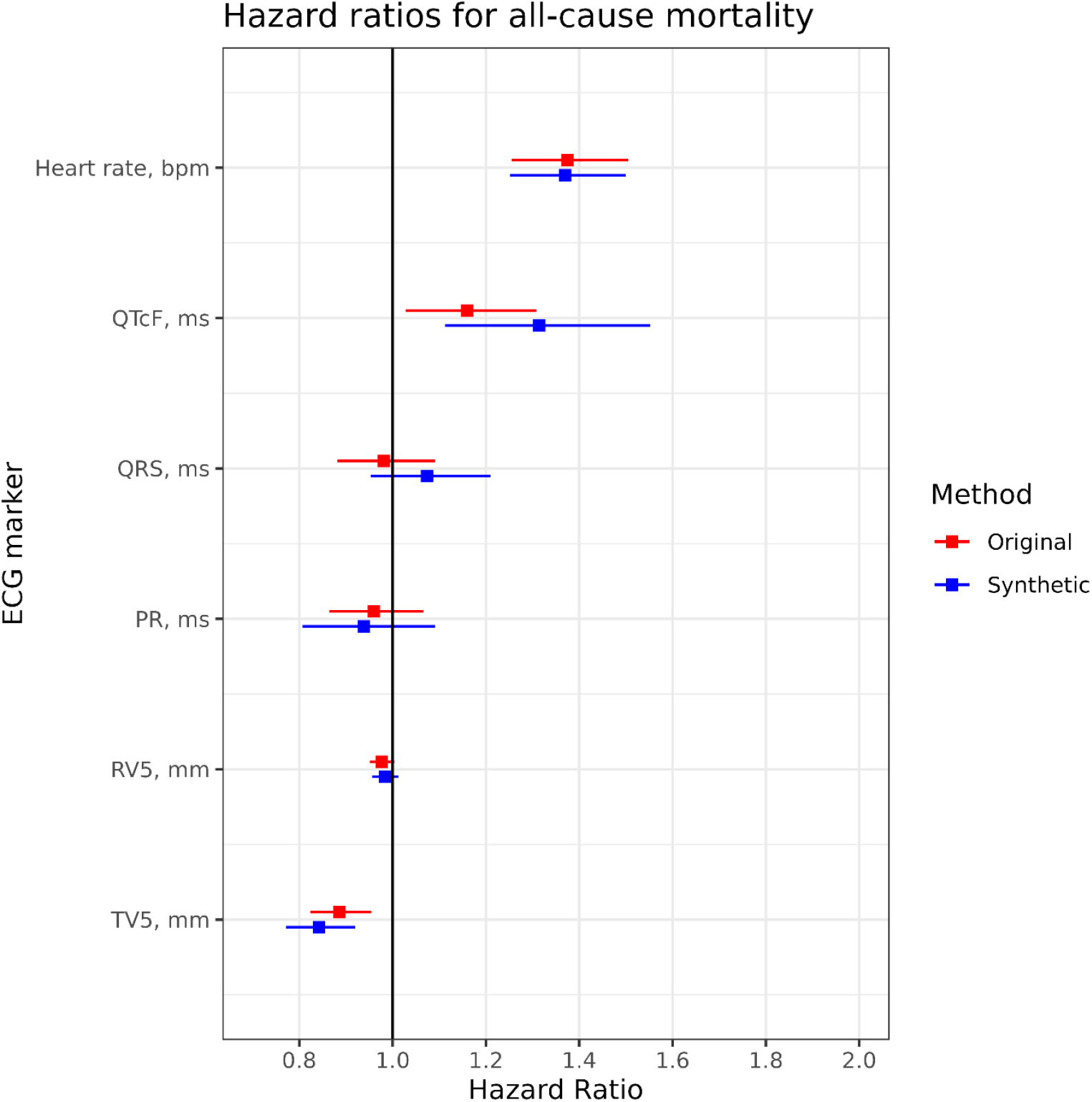
Associations between electrocardiogram (ECG) markers and all-cause mortality. All associations between ECG markers and all-cause mortality were similar in the original and generated populations. Results are individual models adjusted for sex, age, and body mass index (BMI).

## Discussion

We present the first successful tool for synthesizing personalized medical time series, validated with the synthesis of personalized ECGs. The published model requires no additional training or fine-tuning and needs only a single ECG from each individual to be operational.

Previous work demonstrated that the addition of synthetic data in algorithm development results in better detection of myocardial infarction^20^ and atrial fibrillation^21^ and better beat identification.^22^ The authors showed that synthetic data helped overcome class imbalance,^21^ because people with atrial fibrillation are underrepresented in studies despite atrial fibrillation being the most common sustained cardiac arrhythmia.

The value of personalized synthetic ECGs lies in the application of the synthetic ECGs. A lack of metadata precludes current synthetic data methods from being useful for epidemiological studies – cross-sectional studies or time-to-event studies – which is the cornerstone of modern ECG research.^23^ Our work, which combines accurate patient characteristics with ECG synthesis, may allow for the development of risk prediction models based on group sizes that would today be considered too small, because the method allows up-concentration of these groups of patients. Predicting outcomes is the next and much more valuable step compared to diagnostics, because this may help inform and guide patient care. These exciting perspectives should be validated prospectively.

The key challenge in this work was balancing the likeness of synthetic ECGs and source ECGs. On the one hand, creating exact duplicates in practice does not overcome privacy issues and foster data sharing. On the other hand, too dissimilar ECGs do not carry the desired phenotypes of the source ECG. We formulated this elusive balance as the challenge of generating “ECGs, which could have been recorded from a specific individual on another day.”

Given this challenge, it is difficult to assess success because increasing accuracy in synthesizing ECGs will converge on simple copies, which are of no use. However, we were able to assess success by assessing some of the downstream tasks that personalized synthetic ECGs may be used for. Notably, we demonstrated that associations between ECG markers and sex, age, BMI, and even mortality were largely unchanged, enabling epidemiological studies with synthetic ECGs.

The main limitation of our novel method, which used mean squared error as the loss function – was that model training prioritized larger waves over smaller waves. In our case, that was exemplified by the P-wave inaccuracies, because the P wave is by far the smallest wave of the ECG. Our previous non-personal GAN model was also limited by a small P-wave prolongation^10^ (12 ms, ¼ of the smallest square on paper). Thus, the 8 ms prolongation in the current work constitutes an improvement. The location of the P wave in relation to the QRS complex (PR interval), although still personalized to a high degree, showed some evidence of regression to the population mean, leading also to a reduction in the strength of association between PR interval and patient characteristics. However, this shortcoming did not change the association with all-cause mortality significantly.

In this work, we undertook careful validation with an FDA-cleared algorithm (Marquette 12SL) from an industry leader (GE Healthcare), which has been rigorously validated.^15,24^ Currently, common approaches to validation of synthetic ECGs are none, visual inspection, or general metrics such as mean squared error or Frechét distance. However, these methods are not suitable for the validation of medical time series, which require domain-specific validation.^25^ We encourage that this method, when applied to other time series, be validated with proper domain-specific markers.

We are not the first to generate synthetic ECGs, but we were the first to generate personalized synthetic ECGs with a single model. Seminal work on general synthetic ECGs, *ECGSYN*^26^, is hosted on PhysioNet^27^ and produces single-lead ECGs based on three coupled ordinary differential equations. The method was subsequently expanded to create waves separately^28^ and to create 12-lead ECGs, which is the clinical gold standard.^29,30^ Our method is a derivative of a GAN, which has been widely used to make general synthetic ECGs but not personal synthetic ECGs.^10,31,32^

Attempts at personalized ECG generation exist. Golany and Radinsky suggested to train one GAN model per patient, thereby making a general method personalized.^33^ However elegant, the method is infeasible for large datasets. Tang et al. suggested to create synthetic ECGs based on patient-specific photoplethysmograms, but that model also required fine-tuning via an ECG, making the approach less useful because it now requires the acquisition of photoplethysmograms on top of ECGs.^34^ The original *ECGSYN* paper suggested that their method could generate patient-specific (single lead) ECGs,^26^ but to our knowledge that was never validated.

Personalized ECG generation also has potential for training of foundation models for ECG analysis. Current foundation models for ECG are based on contrastive learning, which is implemented by pairing matching ECGs.^35^ This task would be trivial if blanking or noise were not introduced.^35^ We propose that personalized synthetic ECGs be used in the training of foundation models, because the inherent variation by the synthetic method makes the task non-trivial.

In conclusion, we have demonstrated that a novel neural network-based method can generate personalized synthetic ECGs, which retained known associations with all-cause mortality, and which were associated with patient characteristics similarly to the original ECGs.

## Data Availability

We are prohibited by law from sharing the underlying patient data.

## Sources of Funding

JLI is supported by a grant from the Danish Cardiovascular Academy (PD2Y-2023004-DCA). The Danish Cardiovascular Academy is funded by the Novo Nordisk Foundation and the Danish Heart Foundation, grant number NNF20SA0067242. MN is supported by a grant from the Danish Data Science Academy and the Danish Cardiovascular Academy (PhD2024015-DCA-DDSA). The Danish Data Science Academy is funded by the Novo Nordisk Foundation (NNF21SA0069429) and the VILLUM FONDEN (40516). DL is supported by a Novo Nordisk Foundation Young Investigator Awards 2021 (NNF21OC0066480). CE is partly funded from the Laboratory Endowment Fund at Boston Children’s Hospital, USA. GESUS was funded by the Region Zealand Research Foundation, Naestved Hospital Foundation, Naestved Municipality, Johan and Lise Boserup Foundation, TrygFonden, Johannes Fog’s Foundation, Region Zealand, Naestved Hospital, The National Board of Health, The Local Government Denmark Foundation

## Disclosures

None

## Supplementary Material

Supplementary Methods (Instructions on how to use the generative model, Minimal Python code to load and use the model)

Figure S1

## Notes

### Competing Interest Statement

The authors have declared no competing interest.

### Author Declarations

The present study used already collected data from population studies, which had each been approved by the relevant IRB and the use of the data was approved by the Danish Data Protection Agency. The data used in our study was de-identified individual-level data.

